# Pre-endoscopic screening of precancerous lesions in gastric cancer using deep learning

**DOI:** 10.1101/2024.04.08.24305062

**Authors:** Lan Wang, Qian Zhang, Peng Zhang, Bowen Wu, Shiyu Du, Kaiqiang Tang, Shao Li

## Abstract

**Objective:** Given the high cost of endoscopy in gastric cancer (GC) screening, there is an urgent need to explore cost-effective methods for the large-scale prediction of precancerous lesions of gastric cancer (PLGC). We aim to construct a hierarchical artificial intelligence-based multimodal non-invasive method for pre-endoscopic risk screening, to provide tailored recommendations for endoscopy.

**Design:** From December 2022 to December 2023, a large-scale screening study was conducted in Fujian, China. Based on traditional Chinese medicine theory, we simultaneously collected tongue images and inquiry information from 1034 participants, considering the potential of these data for PLGC screening. Then, we introduced inquiry information for the first time, forming a multimodality artificial intelligence model to integrate tongue images and inquiry information for pre-endoscopic screening. Moreover, we validated this approach in another independent external validation cohort, comprising 143 participants from the China-Japan Friendship Hospital.

**Results:** A multimodality artificial intelligence-assisted pre-endoscopic screening model based on tongue images and inquiry information (AITonguequiry) was constructed, adopting a hierarchical prediction strategy, achieving tailored endoscopic recommendations. Validation analysis revealed that the area under the curve (AUC) values of AITonguequiry were 0.74 for PLGC (95% confidence interval (CI) 0.71 to 0.76, p < 0.05) and 0.82 for high-risk PLGC (95% CI 0.82 to 0.83, p < 0.05), which were significantly and robustly better than those of the independent use of either tongue images or inquiry information alone, and also demonstrated superior performance compared to existing screening methodologies. In the independent external verification, the AUC values were 0.69 for PLGC and 0.76 for high-risk PLGC.

**Conclusion:** Our AITonguequiry artificial intelligence model, for the first time, incorporates inquiry information and tongue images, leading to a higher precision and finer-grained pre-endoscopic screening of PLGC. This enhances patient screening efficiency and alleviates patient burden.

## 1 Introduction

According to recent surveys, gastric cancer is the fourth leading cause of cancer-related deaths worldwide and the second in China [1]. There are approximately 480,000 new cases and 370,000 deaths of gastric cancer in China each year, accounting for half of the cases in the world [2]. Gastric cancer is thought to develop from precancerous lesions of gastric cancer (PLGC) (e.g., chronic atrophic gastritis, intestinal metaplasia, or gastric epithelial dysplasia), and the graded screening and diagnosis of natural populations for PLGC is essential to reduce gastric cancer mortality [3–5]. However, the screening and diagnosis of gastric diseases still rely on gastroscopy, but its application is greatly limited because of its invasiveness, high cost, and the need for professional endoscopists [6]. Meanwhile, endoscopic screening is not suitable for large-scale natural populations, especially in rural China. Alternatively, the application of serum markers that are commonly used as screening factors in various gastric cancer risk assessment methods, such as pepsinogen I/II and gastrin-17, has been limited for risk screening in natural populations due to its invasiveness, and the high sensitivity and specificity thresholds [7, 8]. Therefore, there is an urgent need for affordable and non-invasive screening methods suitable for large-scale natural populations to improve diagnostic efficiency and reduce the incidence of gastric cancer.

Early studies have indicated that non-invasive features, including imaging characteristics and clinical phenotypic information, have the potential to predict the occurrence and progression of PLGC. The theory of Traditional Chinese medicine (TCM) suggests that the tongue’s shape, color, size, and coating characteristics detected using tongue images reflect health status and disease severity/progression, and recent studies have shown the potential for tongue surface and color characteristics to assist in the diagnosis of PLGC [9, 10]. For digestive diseases, tongue images characteristics have been found to correlate with gastroscopic observations and predict gastric mucosal health [11, 12]. Additionally, interrogating inquiry information (e.g., living habits, dietary preferences, and physical symptoms) is crucial in understanding the disease and medical history [13]. In this regard, recent studies have built risk prediction models for PLGC before endoscopy using demographics and clinical risk factors, including H. pylori infection, sex, age, race/ethnicity, and smoking status [14]. However, the integration of tongue images and inquiry information for high-precision endoscopic screening of PLGC to facilitate precise endoscopic recommendations has not been studied.

With the rapid development of artificial intelligence (AI) technology, machine learning algorithms based on deep neural networks can accurately analyze diagnostic clinical images, identify therapeutic targets, and process large datasets, which can play a role in screening and diagnosis of a variety of diseases [15–18]. Pan et al. reviewed studies related to AI methods for lung cancer risk prediction, diagnosis, prognosis, and treatment response monitoring [19]. Wang et al. construct an AI-based model of two-dimensional shear wave elastography of the liver and spleen to precisely assess the risk of GEV and high-risk gastroesophageal varices [20]. Ma et al. constructed the deep learning model for screening precancerous lesions of gastric cancer based on tongue images [21]. Li et al. found that both tongue images and the tongue-coating microbiome can be used as tools for the diagnosis of gastric cancer [22]. However, the value of these features in pre-endoscopic PLGC risk screening remains uncertain, and integrating these features based on AI to achieve a more refined PLGC risk screening still poses significant challenges.

In this study, we integrated tongue image data and inquiry data to construct an AI-based multimodal model to assist in pre-endoscopic screening of PLGC. We evaluated the performance of predicting PLGC and high-risk PLGC in a cohort of patients diagnosed with chronic gastritis in Fujian, China, and assessed the superiority of multimodal fusion over single modality. Additionally, we validated the model’s performance in another independent cohort.

## 2 Materials and methods

### 2.1 Design and overview

This research recruited a cohort of patients diagnosed with chronic gastritis from Fujian, China, and the recruitment period spanned from December 2022 to December 2023. Those who volunteered to participate were included in this study. This study was approved by the ethics committee of the Fujian Medical University (Approval number 58 in 2020).

### 2.2 Patient enrollment

One thousand and thirty-four potentially eligible patients were enrolled in this study. The inclusion criteria were as follows: 1) age between 18 and 70 years; 2) have the gastroscopy examination results saved within the past three months, or will undergo gastroscopy examination in the coming three months; 3) no previous diagnosis of cancer; 4) resides locally and is willing to cooperate with doctor’s follow-up; and 5) written informed consent. Patients were excluded for the following reasons: 1) cancer patients; 2) contraindications for endoscopic examination; 3) pregnant or women planning pregnancy, as well as lactating women; 4) cardiovascular, pulmonary, renal, endocrine, neurological, and hematological disorders; 5) mental disorder; and 6). unwilling to participate or poor compliance.

### 2.3 Tongue images and inquiry information

It is recommended that patients adhere to a standardized procedure to acquire high-quality tongue images. Patients are advised to present themselves in natural light conditions during the morning, ensuring an empty stomach. Patients should protrude their tongue from the oral cavity, with particular attention to positioning the tip slightly downward and flattening the surface to ensure the entire tongue body is adequately visualized.

Simultaneously, the healthcare practitioner will engage in a comprehensive traditional Chinese medicine consultation with patients. This consultation encompasses an exploration of demographic details, such as gender, along with an assessment of pertinent lifestyle factors, including a history of smoking and alcohol consumption. Additionally, an inquiry into the patient’s family medical history, dietary habits, and an evaluation of physical symptoms will be conducted. The physical symptoms evaluation involves an assessment of potential discomfort in the stomach and mouth, the patient’s mental state, and their bowel and urinary habits.

### 2.4 Endoscopic evaluation

Two independent gastroenterology experts, each of whom had carried out more than 1000 endoscopies, performed esophagogastroduodenoscopy (EGD) on all patients. The biopsy results were reported as normal, superficial gastritis, chronic atrophic gastritis, intestinal metaplasia, or intraepithelial neoplasia, and a diagnosis was assigned to each participant based on the most severe histological finding in the biopsy. The Helicobacter pylori (Hp) infection status was determined by enzyme-linked immunosorbent assay of plasma IgG [23]. The procedure was conducted up to 3 months before or after the acquisition of images and traditional Chinese medicine inquiry, and the operators were unaware of the results of the tongue examination and inquiry information.

### 2.5 Single-modality deep Learning Risk Prediction Models

#### 2.5.1 Tongue images deep learning risk prediction (Single-Tongue) model

This section proposes Single-Tongue, a new diagnostic approach based on single-modality deep learning using tongue images to predict PLGC and high-risk PLGC. We applied the Segment Anything Models to segment tongue images to extract the features of the effective area and avoid the influence of irrelevant edge noise information.

All patients were randomly divided into training and validation cohorts. The training cohort was utilized to train a deep neural network designed for this study. The performance of the trained model was evaluated through its application to the validation cohort. In order to extract the features from the tongue images, we employed a pre-trained ResNet framework that had been previously trained on the ImageNet dataset [24]. Distinct from convolutional neural networks (CNNs), ResNet tackles the issues of vanishing gradients and network degradation by introducing direct skip connections within the network, which retain a certain proportion of the output from the previous network layer. Data augmentation techniques such as random cropping, flipping, and rotation were applied to all image data to mitigate overfitting. The images were passed through ResNet during training, specifically through the bottleneck and residual units. After passing through 12 bottleneck layers, an adaptive average pooling operator was used to obtain image features, which were then flattened to a size of 2048*1. The final classification results were generated through a softmax layer. In the single-modality experiment, two binary classification networks were trained to determine the presence or absence of PLGC or high-risk PLGC.

#### 2.5.2 Inquiry information deep learning risk prediction (Single-Inquiry) model

The inquiry information encompasses variables such as sex, age, and the individuals’ history of smoking and alcohol consumption. Furthermore, it incorporates the family medical history, dietary habits, and physical symptoms of the patients, including discomfort in the stomach and mouth and their mental state. We employed a segregation approach by categorizing the features into numerical and factor types to enhance the effectiveness of utilizing the inquiry information. The numerical features were subjected to min-max normalization, scaling them between 0 and 1. On the other hand, the factor features were transformed into numeric vectors using keyword-based encoding techniques.

After feature filtering and mapping, the inquiry information was input into a multilayer perceptron to obtain corresponding feature vectors. Then, two binary classification networks were trained to determine the presence or absence of PLGC or high-risk PLGC.

### 2.6 Multimodality deep learning risk prediction (AITonguequiry) model

Medical data are frequently multimodal. For instance, both tongue images and inquiry information encompass details associated with PLGC. Consequently, in this section, we integrated these two modalities with an attentional mechanism.

In the multimodality experiment, similar to the single-modality experiment, we trained two binary classification networks to determine the presence or absence of PLGC or high-risk PLGC. We employed the dropout method to eliminate a certain proportion of model parameters. Subsequently, we utilized the feature embedding method to align the feature vectors of tongue images and inquiry information for comprehensive patient information utilization.

### 2.7 Statistical analysis

The prediction results were validated by quantitative indexes, including sensitivity, specificity, positive predictive value and negative predictive value. The chi-squared test and t test were used to determine whether there was any significant difference in patient characteristics. The area under the receiver operating characteristic (ROC) curve (AUC) was used to estimate the probability that the model would produce a correct prediction. The DeLong test was used to test whether there was a significant difference in risk prediction between AITonguequiry and other methods.

## 3 Results

### 3.1 Patient characteristics

In this research, a cohort of 1034 participants was recruited in Fujian, China, and the recruitment period spanned from December 2022 to December 2023. Among these patients, NPLGC was documented in 855 (82.61%) patients, and PLGC was documented in 180 (17.39%) patients. Among PLGC, low-risk PLGC and high-risk PLGC account for 346(65.90%) and 179(34.10%), respectively. After randomization of these patients, 828 patients were assigned to the training cohort. The other 207 patients composed the validation cohort.

The baseline characteristics of the study population are summarized in Table 1 and Table 2. Overall, the average age of patients with PLGC was 62, and the standard deviation was 7. In terms of gender, the proportion of males with PLGC (180[34.29%]), and females with PLGC (345[65.71%]). Between the training and validation cohorts, there were no significant differences in any of the baseline characteristics (p>0.05) or in the distribution of patients between NPLGC and PLGC.

**Table 1.** Baseline characteristics of the study cohort: NPLGC and PLGC.

| Characteristic | Training and validation dataset<sup>1</sup> |  | p-value<sup>2</sup> |
| --- | --- | --- | --- |
|  | NPLGC, N = 509 | PLGC, N = 525 |  |
| Age (Years) | 59 (9) | 61 (8) | <0.001 |
| Sex (Female/Male) | 343 /166 | 345 /180 | 0.57 |
| Family history (Yes/No) | 39/470 | 47/478 | 0.45 |
| Drinking (Yes/No) | 136/373 | 144/381 | 0.80 |
| Smoking (Yes/No) | 141/368 | 153/372 | 0.61 |
| Hp (Positive/Negative/Unknown) | 14/31/464 | 19/32/474 | 0.73 |

<sup>1</sup>Mean (SD); n (%)
<sup>2</sup>Welch Two Sample t-test; Pearson's Chi-squared test; Fisher's exact test
PLGC, precursor lesions of gastric cancer; NPLGC, Non-precursor lesions of gastric cancer; Hp, helicobacter pylori.

**Table 2.**
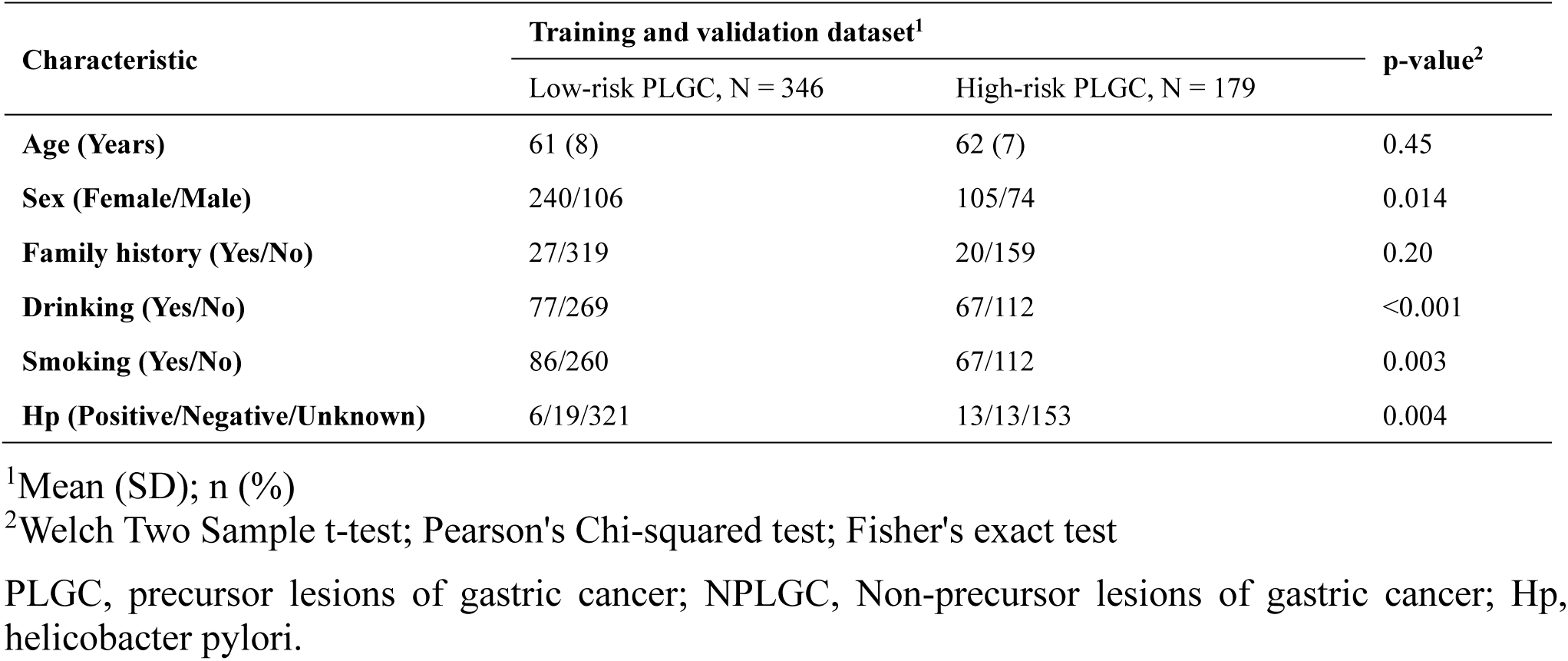
Baseline characteristics of the study cohort: low-risk PLGC and high-risk PLGC.

| Characteristic | Training and validation dataset<sup>1</sup> |  | p-value<sup>2</sup> |
| --- | --- | --- | --- |
|  | Low-risk PLGC, N = 346 | High-risk PLGC, N = 179 |  |
| Age (Years) | 61 (8) | 62 (7) | 0.45 |
| Sex (Female/Male) | 240/106 | 105/74 | 0.014 |
| Family history (Yes/No) | 27/319 | 20/159 | 0.20 |
| Drinking (Yes/No) | 77/269 | 67/112 | <0.001 |
| Smoking (Yes/No) | 86/260 | 67/112 | 0.003 |
| Hp (Positive/Negative/Unknown) | 6/19/321 | 13/13/153 | 0.004 |

<sup>1</sup>Mean (SD); n (%)
<sup>2</sup>Welch Two Sample t-test; Pearson's Chi-squared test; Fisher's exact test
PLGC, precursor lesions of gastric cancer; NPLGC, Non-precursor lesions of gastric cancer; Hp, helicobacter pylori.

### 3.2 Construction of the AITonguequiry model

We build a deep learning risk prediction model based on tongue images and inquiry information (AITonguequiry) from 1034 patients to evaluate their potential in the grading screening and diagnosis of PLGC. The AITonguequiry flow chart is shown in Figure 1. As shown in Figure 1a, the study cohort and the diagnostic model were designed to assess the risk of PLGC and high-risk PLGC based on tongue images and inquiry information. The detailed multimodality model is shown in Figure 1b. The patients were categorized into two groups: Non-PLGC (NPLGC) and PLGC, and the PLGC group was further divided into low-risk PLGC (chronic atrophic gastritis) and high-risk PLGC (intestinal metaplasia or gastric epithelial dysplasia) stages. We advocate for patients predicted with PLGC to undergo endoscopic examination, with a concurrently recommending prompt endoscopic examination for those predicted with high-risk PLGC.

**Figure 1.**
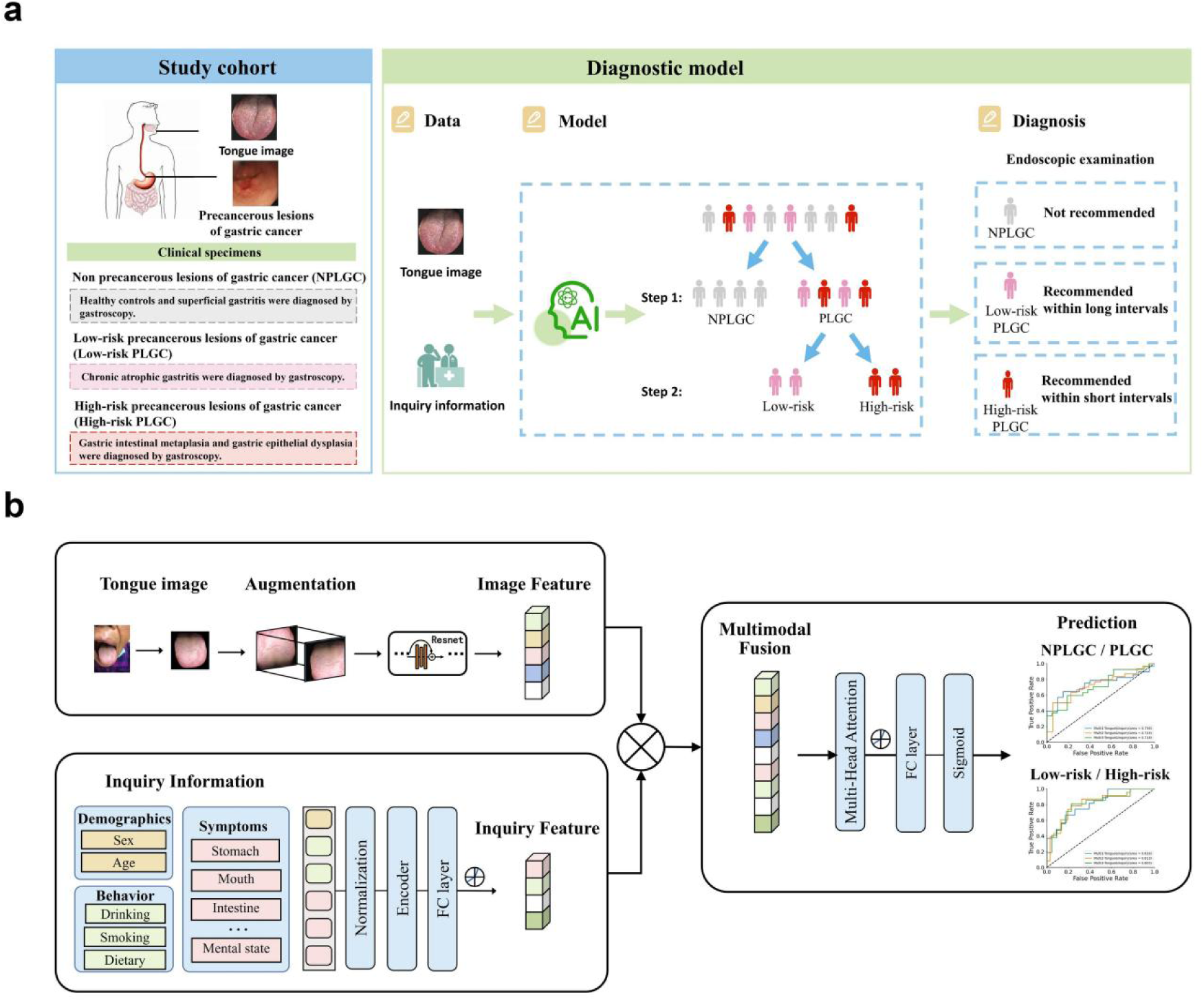
AITonguequiry flow chart. (a) The study cohort and the diagnostic model. (b) A deep learning–based multimodality classification model was developed to assess the risk of PLGC and high-risk PLGC based on the tongue images and inquiry information. PLGC, precursor lesions of gastric cancer; NPLGC, Non-precursor lesions of gastric cancer.

### 3.3 Comparison of the AITonguequiry model, single modality models and baseline characteristics

We chose Single-Tongue, Single-Inquiry and baseline characteristics to identify the presence or absence of PLGC and high-risk PLGC.

The selection of baseline characteristics is based on individuals aged ≥ 45 who meet any of the following criteria, which are indicative of a high-risk profile for gastric cancer: 1) Long-term residence in high-incidence areas of gastric cancer; 2) Hp infection; 3) History of chronic atrophic gastritis, gastric ulcer, gastric polyp, residual stomach after surgery, hypertrophic gastritis, pernicious anemia, or other precancerous diseases of the stomach; 4) First-degree relatives with a history of gastric cancer; 5) Presence of other high-risk factors for gastric cancer such as high salt intake, pickled diet, smoking, and heavy alcohol consumption [25–28]. Since our screening is conducted in high-risk areas, we ignore the first criterion.

In identifying the presence or absence of PLGC, AITonguequiry demonstrated statistically higher AUCs than Single-Tongue, Single-Inquiry and baseline characteristics (p < 0.05) (Figure 2a, Table 3). Impressively, AITonguequiry had an AUC of 0.736 for the diagnosis of PLGC, which was higher than other methods (Figure 2a, all p <0.05). The sensitivity and specificity analyses also demonstrated that AITonguequiry universally outperformed the Single-Tongue, Single-Inquiry and baseline characteristics for assessing PLGC and high-risk PLGC (Table 3).

**Figure 2.**
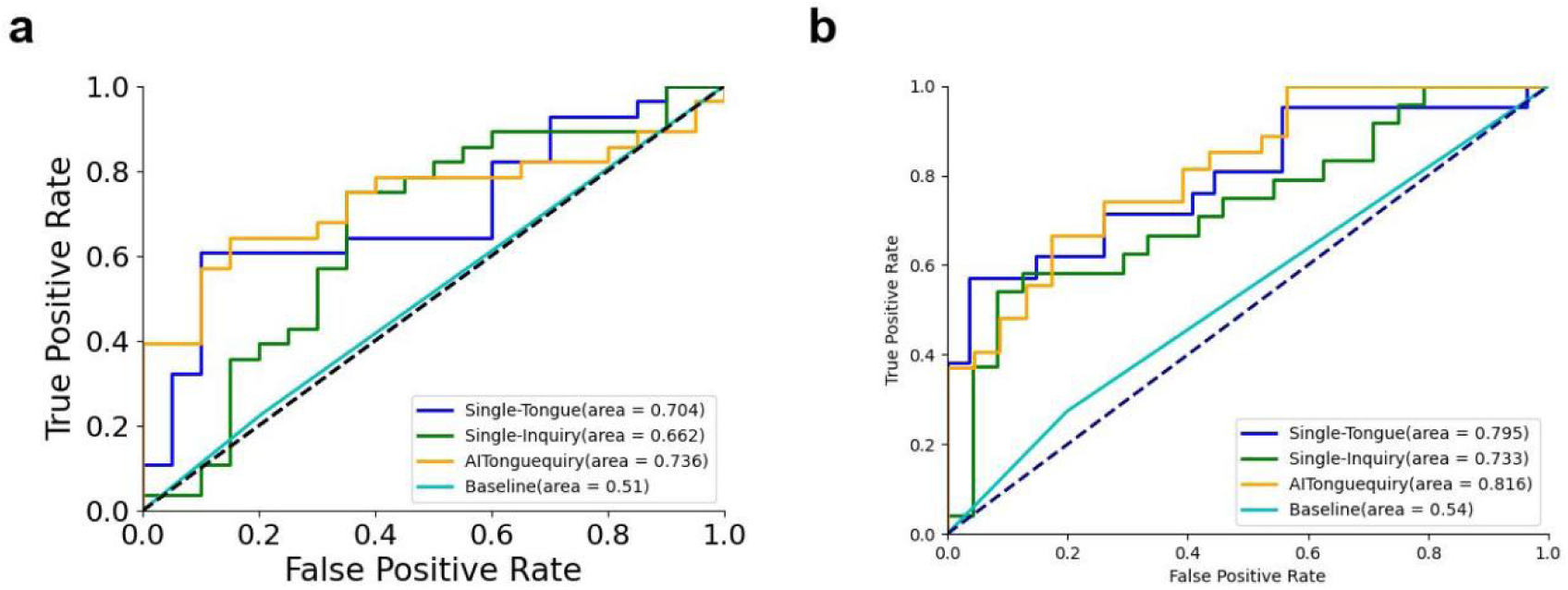
Comparison of ROC curves between different methods for classifying the presence or absence of PLGC and high-risk PLGC in the validation cohorts. (a) Presence or absence of NPLGC/PLGC in the validation cohorts. (b) Presence or absence of low-risk PLGC/high-risk PLGC in validation cohorts. PLGC, precursor lesions of gastric cancer; NPLGC, Non-precursor lesions of gastric cancer; Single-Tongue, single-modality deep learning risk prediction with tongue images; Single-Inquiry, single-modality deep learning risk prediction with inquiry information; AITonguequiry, multimodality deep learning risk prediction with tongue images and inquiry information.

**Table 3.** Diagnostic performance of AITonguequiry for the assessment of NPLGC/PLGC and low-risk PLGC/high-risk PLGC in the validation cohorts.

|  |  | AUC | Specificity (%) | Sensitivity (%) | PPV (%) | NPV (%) |
| --- | --- | --- | --- | --- | --- | --- |
| Step 1: | Single-Tongue | 0.70* | 54.55 | 86.67 | 46.43 | 90.00 |
| NPLGC |  | (0.68 to 0.73) | (53.14 to 56.02) | (84.23 to 89.18) | (43.65 to 49.31) | (87.99 to 91.96) |
| /PLGC | Single-Inquiry | 0.66* | 62.50 | 68.75 | 78.57 | 50.00 |
|  |  | (0.64 to 0.69) | (59.63 to 65.58) | (67.13 to 70.32) | (76.33 to 80.96) | (46.58 to 53.42) |
|  | AITonguequiry | 0.74 | 66.67 | 73.33 | 78.57 | 60.00 |
|  |  | (0.71 to 0.76) | (63.94 to 69.43) | (71.59 to 75.04) | (76.16 to 80.87) | (56.66 to 63.27) |
| Step 2: |  |  |  |  |  |  |
| Low-risk | Single-Tongue | 0.79* | 90.00 | 52.63 | 95.24 | 33.33 |
| PLGC |  | (0.78 to 0.82) | (86.97 to 92.58) | (51.54 to 53.69) | (93.60 to 96.57) | (30.58 to 36.00) |
| /High-risk | Single-Inquiry | 0.73* | 64.00 | 65.22 | 62.50 | 66.67 |
| PLGC |  | (0.71 to 0.75) | (61.91 to 66.18) | (63.03 to 67.39) | (59.50 to 65.60) | (63.80 to 69.50) |
|  | AITonguequiry | 0.82 | 66.67 | 78.26 | 66.67 | 78.26 |
|  |  | (0.80 to 0.83) | (64.61 to 68.66) | (76.09 to 80.44) | (63.79 to 69.44) | (75.52 to 80.85) |

The AUC of AITonguequiry was statistically compared with the AUCs of Single-Tongue and Single-Inquiry (\*P<0.05; \*\*P<0.01). AUC, area under the receiver operating characteristic curve; PLGC, precursor lesions of gastric cancer; NPLGC, Non-precursor lesions of gastric cancer; Single-Tongue, single-modality deep learning risk prediction with tongue images; Single-Inquiry, single-modality deep learning risk prediction with inquiry information; AITonguequiry, multimodality deep learning risk prediction with tongue images and inquiry information; PPV, positive predictive value; NPV, negative predictive value.

In identifying the presence or absence of high-risk PLGC, AITonguequiry demonstrated statistically higher AUCs than Single-Tongue, Single-Inquiry and baseline characteristics (p < 0.05) (Figure 2b, Table 3). Impressively, AITonguequiry had an AUC of 0.816 for the diagnosis of high-risk PLGC, which was higher than other methods (Figure 2b, all p <0.05). The sensitivity and specificity analyses also demonstrated that AITonguequiry universally outperformed the Single-Tongue, Single-Inquiry and baseline characteristics for assessing PLGC and high-risk PLGC (Table 3).

### 3.4 Evaluation of the diagnostic robustness of the AITonguequiry model

All patients were randomly divided into training and validation cohorts. Simultaneously, all the images of each patient were allocated to the cohort corresponding to that patient, ensuring that there was no simultaneous presence of different images from the same patient in both cohorts. In either the validation cohort, the results in the three ROC curves always overlapped each other (Figure 3), and no significant differences were found (all p>0.05, Table 4). These results revealed that AITonguequiry demonstrated robust and consistent performances regardless of the data from which medical centers, as long as the number of enrolled patients in different training cohorts was fairly constant.

**Figure 3.**
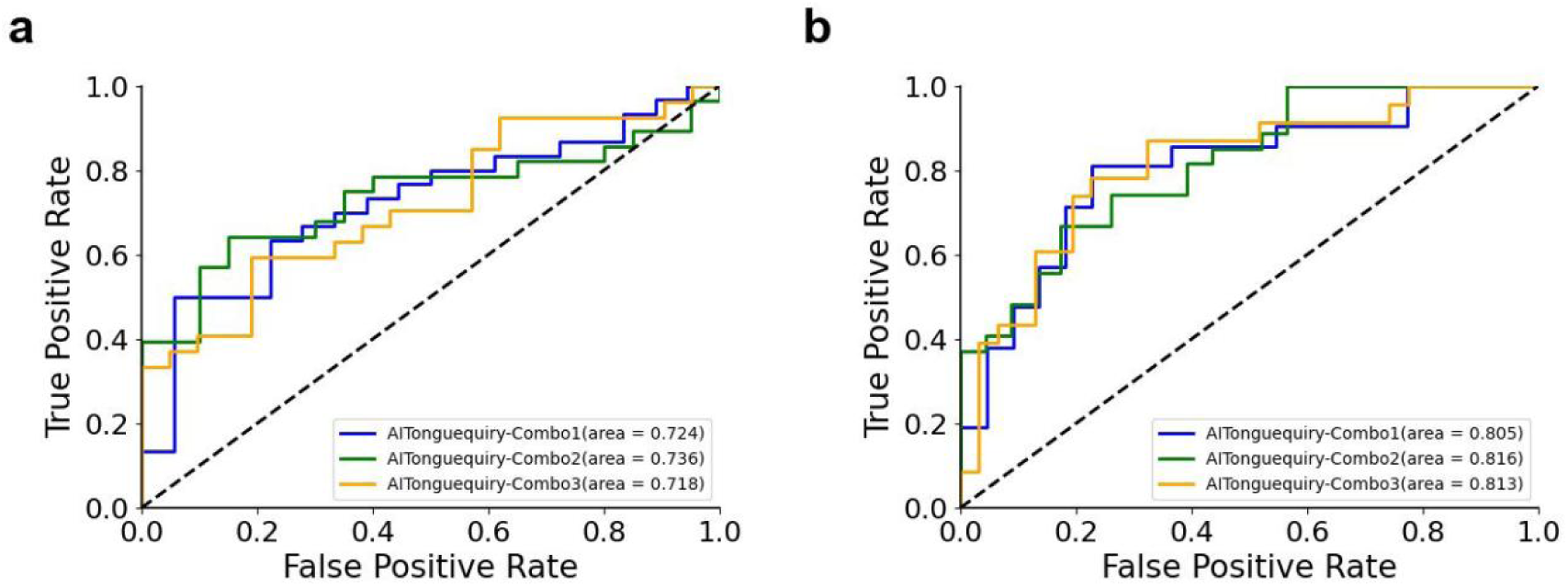
Comparison of receiver operating characteristic (ROC) curves among different combinations using AITonguequiry. (a) Presence or absence of NPLGC/PLGC in the validation cohorts. (b) Presence or absence of low-risk PLGC/high-risk PLGC in validation cohorts. PLGC, precursor lesions of gastric cancer; NPLGC, Non-precursor lesions of gastric cancer; AITonguequiry, multimodality deep learning risk prediction with tongue images and inquiry information.

**Table 4.** Diagnostic robustness of AITonguequiry for the assessment of NPLGC/PLGC and low-risk PLGC/high-risk PLGC in the validation cohorts.

|  | AUC | Specificity (%) | Sensitivity (%) | PPV (%) | NPV (%) |
| --- | --- | --- | --- | --- | --- |
| Step1: |  |  |  |  |  |
| NPLGC/PLGC |  |  |  |  |  |
| Combo 1 | 0.74 | 66.67 | 73.33 | 78.57 | 60.00 |
|  | (0.71 to 0.76) | (63.91 to 69.44) | (71.62 to 75.06) | (76.16 to 80.88) | (56.90 to 63.27) |
| Combo 2 | 0.72 | 54.55 | 76.92 | 66.67 | 66.67 |
|  | (0.70 to 0.75) | (52.21 to 56.88) | (75.00 to 78.79) | (64.00 to 69.20) | (63.33 to 70.00) |
| Combo 3 | 0.72 | 60.00 | 67.86 | 70.37 | 57.14 |
|  | (0.69 to 0.74) | (57.32 to 62.52) | (65.96 to 69.73) | (67.73 to 73.07) | (53.83 to 60.46) |
| Step2: |  |  |  |  |  |
| Low-risk PLGC |  |  |  |  |  |
| /High-risk PLGC |  |  |  |  |  |
| Combo 1 | 0.82 | 66.67 | 78.26 | 66.67 | 78.26 |
|  | (0.80 to 0.83) | (64.63 to 68.66) | (75.95 to 80.51) | (63.80 to 69.35) | (75.30 to 80.96) |
| Combo 2 | 0.81 | 73.91 | 75.00 | 71.43 | 77.27 |
|  | (0.79 to 0.82) | (72.00 to 76.09) | (72.80 to 77.17) | (68.65 to 74.39) | (74.78 to 79.77) |
| Combo 3 | 0.81 | 86.36 | 62.50 | 86.96 | 61.29 |
|  | (0.79 to 0.83) | (84.22 to 88.53) | (60.66 to 64.32) | (84.61 to 89.31) | (58.45 to 64.11) |

Statistical values are presented with the 95% CIs when applicable. AUCs obtained with three different combinations of patients were statistically compared with each other in each classification and each cohort (\*P<0.05; \*\*P<0.01). AUC, area under the receiver operating characteristic curve; PLGC, precursor lesions of gastric cancer; Combo, combination of patients for training and validation cohorts; PPV, positive predictive value; NPV, negative predictive value.

### 3.5 Independent external validation of the AITonguequiry model

Moreover, 143 participants were recruited for independent external validation from the China-Japan Friendship Hospital. In the independent validation cohort, the AUC of PLGC was 0.69, and the AUC of high-risk PLGC was 0.77 (Figure 4, Table 5). These results demonstrate the effectiveness of the AITonguequiry model in independent external validation.

**Figure 4.**
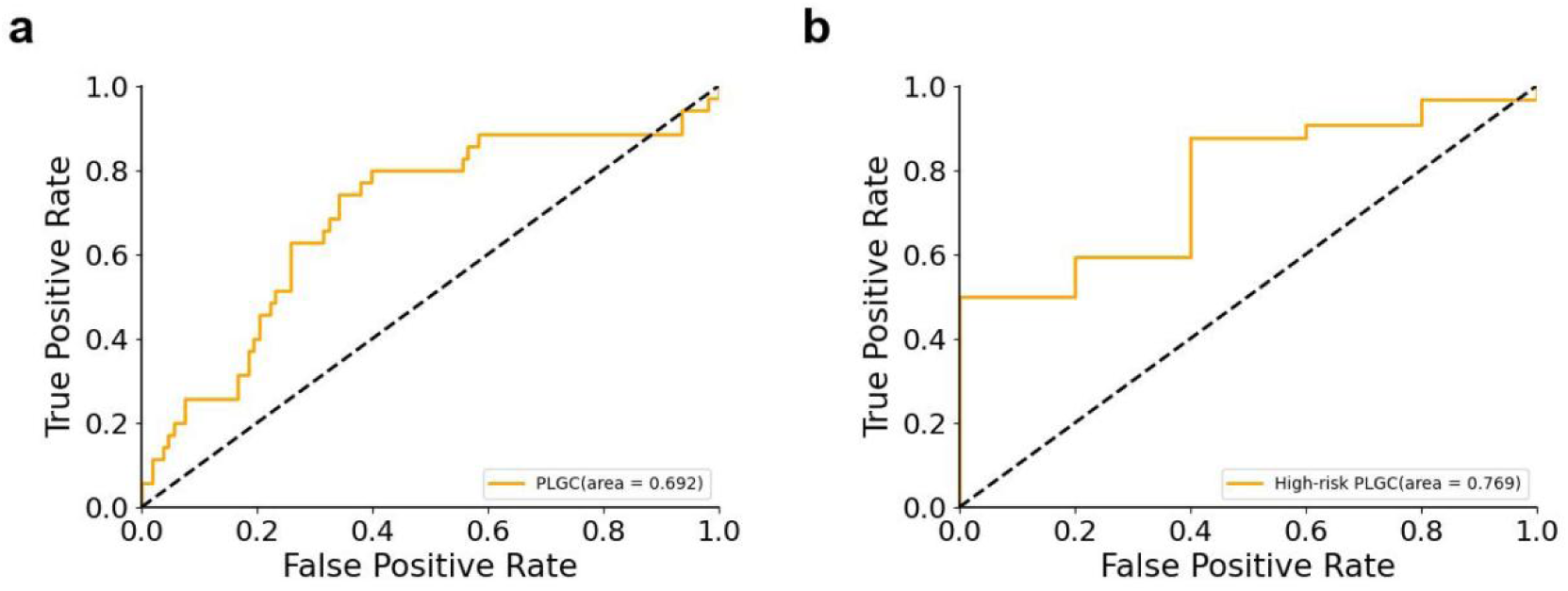
Receiver operating characteristic (ROC) curves during the external validation using Single-Tongue. Presence or absence of NPLGC/PLGC in the external validation cohorts and presence or absence of low-risk PLGC/high-risk PLGC in the external validation cohorts. PLGC, precursor lesions of gastric cancer; NPLGC, Non-precursor lesions of gastric cancer.

**Table 5.** Diagnostic performance of Single-Tongue for the assessment of NPLGC/PLGC and low-risk PLGC/high-risk PLGC in the external validation cohorts.

|  | AUC | Specificity (%) | Sensitivity (%) | PPV (%) | NPV (%) |
| --- | --- | --- | --- | --- | --- |
| Step 1: |  |  |  |  |  |
| NPLGC/PLGC | 0.69<br>(0.66 to 0.72) | 80.56<br>(79.36 to 81.73) | 40.00<br>(36.36 to 43.56) | 40.00<br>(35.58 to 44.18) | 80.56<br>(78.61 to 82.52) |
| Step 2: |  |  |  |  |  |
| Low-risk PLGC | 0.77 | 33.33 | 88.24 | 93.75 | 20.00 |
| /High-risk PLGC | (0.74 to 0.79) | (26.49 to 40.00) | (87.61 to 88.90) | (92.60 to 94.85) | (15.19 to 24.81) |

AUC, area under the receiver operating characteristic curve; PLGC, precursor lesions of gastric cancer; NPLGC, Non-precursor lesions of gastric cancer; Single-Tongue, single-modality deep learning risk prediction with tongue images; Single-Inquiry, single-modality deep learning risk prediction with inquiry information; AITonguequiry, multimodality deep learning risk prediction with tongue images and inquiry information; PPV, positive predictive value; NPV, negative predictive value.

## 4 Discussion

Given the significant burden of GC in China and globally, it becomes imperative to adopt cost-effective approaches for large-scale screening of PLGC risk prediction in the natural population. While gastroscopy and pathological tests serve as the gold standard for diagnosing gastric diseases, they are not suitable for widespread application in the natural population. To address this pressing issue, there is an urgent need to develop noninvasive and effective screening and diagnostic methods to provide tailored recommendations for endoscopy. Deep neural network technology offers a promising avenue for advancing healthcare systems by providing heightened accuracy and computational power, thereby playing an increasingly vital role in disease risk prediction. In our study, a multimodality AI-assisted pre-endoscopic screening model based on tongue images and inquiry information (AITonguequiry) was constructed for the first time, adopting a hierarchical prediction strategy, achieving tailored endoscopic recommendations.

In this study, the diagnostic accuracy of AITonguequiry was significantly better than that of Single-Tongue or Single-Inquiry in assessing PLGC and high-risk PLGC. In the evaluation of PLGC, the AUC in the validation cohort was 0.74 (Figure 2a, Table 3). Thus, AITonguequiry was effective for the assessment of PLGC. In the evaluation of high-risk PLGC, the AUC in the verification cohort was 0.82, showing that AITonguequiry was effective for the assessment of high-risk PLGC (Figure 2b, Table 3). The above analysis revealed that the multimodality models were effective in discriminating patients with PLGC from participants without PLGC, and could also effectively differentiate patients with high-risk PLGC from those with low-risk PLGC. Thus, the fusion model of tongue images and inquiry information further improved the diagnostic value (Figure 2, Table 3).

The risk prediction of PLGC using tongue images and inquiry information can serve as an effective, noninvasive auxiliary diagnostic method that can support primary healthcare systems worldwide. With the recent advancements in deep neural networks, significant progress has been made in standardizing tongue images and inquiry information risk prediction, especially in TCM. Many results have been achieved in tongue image preprocessing, tongue detection, segmentation, feature extraction and tongue analysis [28]. Xu et al. developed a multi-task joint learning model to segment and classify tongue images using deep neural networks, which can optimally extract tongue image features [29]. Li et al. pioneered the use of both tongue images and the tongue-coating microbiome as diagnostic tools for gastric cancer [22]. Similarly, Ma et al. constructed the first deep learning model for screening precancerous lesions of gastric cancer based on tongue images [21]. To our knowledge, AITonguequiry is the first simplified, novel, AI-based image processing tool that can accurately and noninvasively identify PLGC and high-risk PLGC. Moreover, our AITonguequiry multimodal model, for the first time, incorporates inquiry information and employs a hierarchical prediction strategy, resulting in more refined endoscopic recommendations. We advocate for patients predicted with PLGC to undergo endoscopic examination, with a concurrently recommending prompt endoscopic examination for those predicted with high-risk PLGC. With the AITonguequiry, the operator only needs to carry out the daily data acquisition workflow to analyze the critical information automatically, making this model very convenient for clinical applications, which enhances patient screening efficiency and alleviates patient burden.

According to the principles of TCM, tongue manifestation serves as a crucial indication of “collaterals” [30]. It plays a significant role in various disorders such as gastric diseases [21], rheumatic conditions [31], hepatic disorders [32], and tumors [10], among others. Tongue color, thickness, moisture, and other changes, as well as the rich network of vessels in the tongue area [30], to a certain extent, can reflect the pathological changes in the spleen, stomach, internal organs, joints, and many other parts. Tongue diagnosis and inquiry are integral components of the four diagnostic methods in TCM. The characteristics of the tongue’s shape, color, size, and coating, as detected through tongue images, can reflect an individual’s health status and the severity/progression of diseases [33]. In addition, inquiry information, including demographics, behavior, and physical symptoms, plays a crucial role in understanding the disease and medical history. In this study, we discovered that useful features for predicting PLGC can be extracted from tongue images using a deep learning model. This finding indicates that the evaluation of human health based on tongue characteristics, as proposed by TCM theory, has scientific grounds. Furthermore, we found that the deep-learning model can extract informative features for PLGC prediction from inquiry information, suggesting that the evaluation of human health based on demographic, behavioral, and physical symptom information in inquiries, as guided by TCM theory, is scientifically supported. We believe that with the widespread application of AI and deep learning methods, tongue images and inquiry information can potentially become cost-effective, non-invasive and acceptable approaches for predicting and screening PLGC, which will also lead to significant socio-economic impacts.

Nevertheless, our research has some limitations. One limitation of our study is the restricted number of patients included. For future research, it will be essential to involve a larger screening population to improve the training of the deep learning model. Additionally, there is potential to expand by creating a semantic dataset of tongue images and establishing a multimodal large language model for PLGC prediction, offering personalized predictions and detailed explanations for clinicians and patients. These areas merit further exploration in the future.

In conclusion, this study introduces a hierarchical AI-based multimodal non-invasive method for pre-endoscopic risk screening. All these findings indicate that AITonguequiry is a non-invasive method for predicting and assessing PLGC and high-risk PLGC, showcasing its strong performance in graded screening and diagnosis. Our method has a good potential for widespread clinical use, and further studies in larger patient populations are needed. We will further promote the application of AITonguequiry in PLGC risk prediction and provide tailored recommendations for endoscopy to improve the diagnosis rate, especially high-risk PLGC, in the natural population. Moreover, this study provides scientific support for the theory of tongue images and inquiry information diagnosis in TCM.

## Author contributions

Study conception and design: Shao Li, Peng Zhang, Lan Wang, Qian Zhang and Kaiqiang Tang; Data collection and analysis: Shao Li, Lan Wang, Qian Zhang, Bowen Wu, Shiyu Du and Kaiqiang Tang; The first draft of the manuscript: Lan Wang, Kaiqiang Tang, Qian Zhang; Commented on previous versions of the manuscript: Shao Li, Peng Zhang. All authors read and approved the final manuscript.

## Compliance with Ethical Requirements

## (2) Informed Consent in Studies with Human Subjects

All procedures followed were in accordance with the ethical standards of the responsible committee on human experimentation (institutional and national) and with the Helsinki Declaration of 1975, as revised in 2008 (5). Informed consent was obtained from all patients for being included in the study.

## (3) Data availability

The data of individual deidentified participants will not be shared, but it is available upon request via.

